# Serial Interval Distribution of SARS-CoV-2 Infection in Brazil

**DOI:** 10.1101/2020.06.09.20127043

**Authors:** Carlos A. Prete, Lewis Buss, Amy Dighe, Victor Bertollo Porto, Darlan da Silva Candido, Fábio Ghilardi, Oliver G. Pybus, Wanderson K. de Oliveira, Júlio H. R. Croda, Ester C. Sabino, Nuno Rodrigues Faria, Christl A. Donnelly, Vítor H. Nascimento

## Abstract

Using 65 transmission pairs of SARS-CoV-2 reported to the Brazilian Ministry of Health we estimate the mean and standard deviation for the serial interval to be 2.97 and 3.29 days respectively. We also present a model for the serial interval probability distribution using only two parameters.

## Text

The novel severe acute respiratory syndrome coronavirus type 2 (SARS-CoV-2) emerged in China in December 2019 [1] and was declared a pandemic by the World Health Organization on 11 March 2020. Current assessments of SARS-CoV-2 transmission dynamics rely on accurate estimates of key epidemiological parameters, including the serial interval, which can be defined as the time between symptom onset of the source and the onset of symptoms of the recipient [2]. Moreover, serial interval estimations can help disentangling the duration of pre-symptomatic transmission upon infection. When the serial interval is greater than the incubation period, an epidemic is characterized by symptomatic transmission. However, existence of pre-symptomatic transmission can be inferred when the serial interval is shorter than the incubation period.

We estimate the serial interval of SARS-CoV-2 from 65 infector-infectee pairs from Brazil. Data was provided by the Brazilian Ministry of Health following ethical approval (CONEP protocol number 30127020.0.0000.0068). All confirmed cases of SARS-CoV-2 infection notified on the REDCap system between 25^th^ February and 19^th^ March were analyzed. We received data on infector and infectee pairs by the Brazilian Ministry of Health, and we measured the serial intervals by computing the difference between the dates of symptom onset of each pair.

The median serial interval in our data was estimated at 3 (standard deviation = 3.29) days. We identified 4 (6.15% of 65) negative serial intervals (symptom onset in the infectee preceded the infector) and seven (10.77% of 65) zero-valued serial intervals (symptom onset on the same day for the infector-infectee pair). It is common to censor observed serial intervals to include only positive values; however, there is no strong theoretical justification for this. As such, we fit a serial interval probability distribution that allows for negative values by modelling the serial interval as:

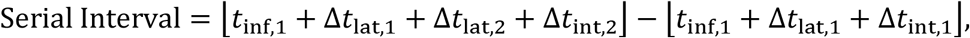

where ⌊⋅⌋ is the floor operator, *t*_inf,1_ is a random variable uniformly distributed in the interval [0,1], (Δ*t*_lat,1,_Δ*t*_lat,2_)are independent and identically distributed (i.i.d.) chi-squared random variables of mean 3.03days and (Δ*t*_int,1_,Δ*t*_int,2_)are i.i.d. chi-squared random variables of mean 0.95days. See the **Supplemental Material** for theoretical motivation, methods and sensitivity analysis.

Figure 1 shows the comparison between the observed and modelled serial intervals as well as the means ±standard deviations for a number of serial interval probability distributions presented in the literature [3–8]. Our proposed model for the serial interval probability distribution approximates well the serial intervals measured from data.

**Figure 1.**
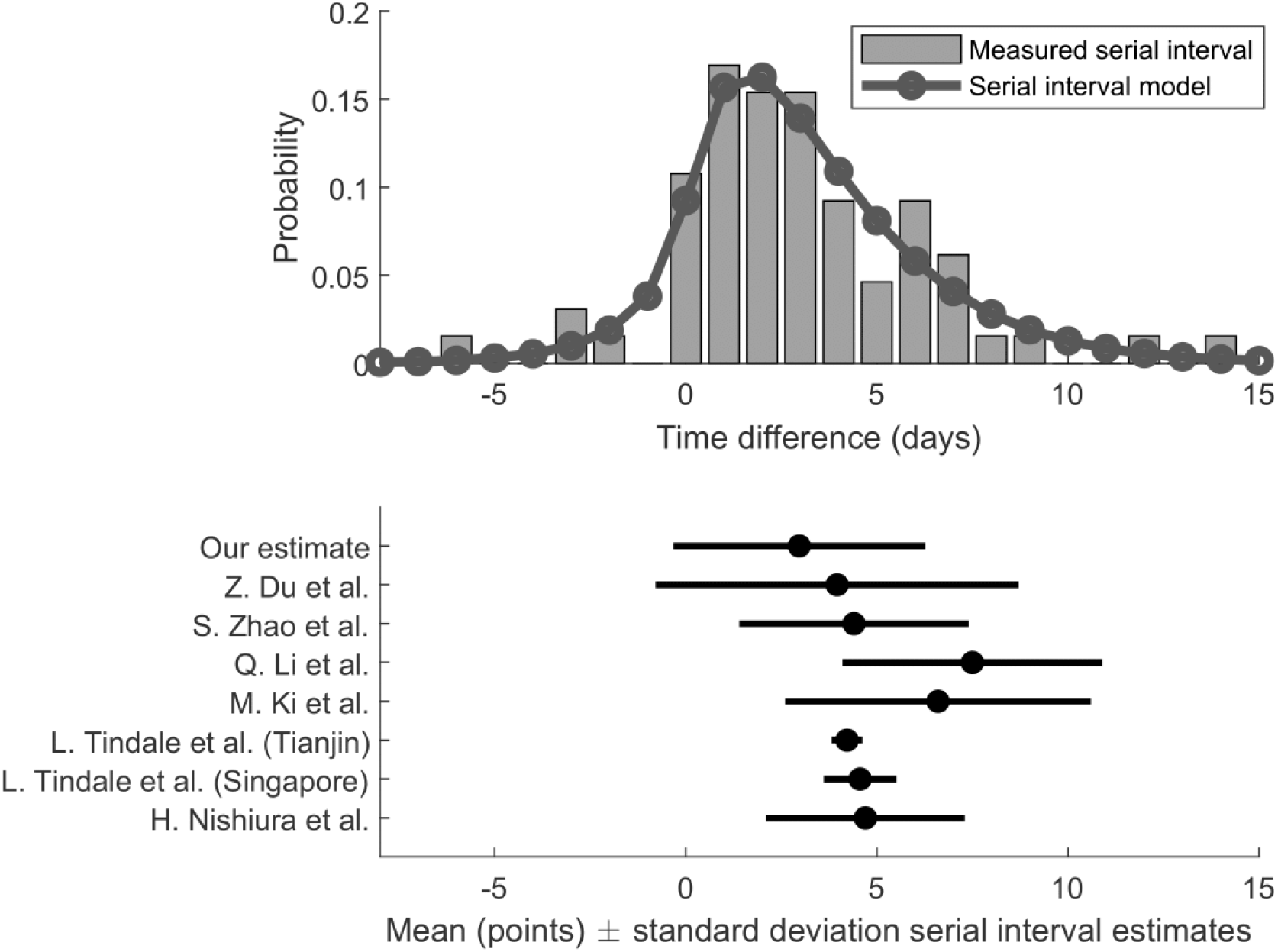
Serial interval estimation from Brazilian SARS-CoV-2 confirmed cases compared to the modelled distribution. Lower panel shows the mean ± standard deviation serial interval estimates from the current literature.

We provide two additional analyses of the data in the Supplemental Material: i) Du et al. [3] took an alternative approach fitting a normal distribution to 468 publicly reported serial intervals in which 12% had negative values. For this reason, we fit a normal distribution to the observed serial intervals and compare it to the distribution presented in [3], obtaining a lower mean and variance. ii.) We censor the data to include only positive values, obtaining a distribution of mean 3.83 days that is best fit by a lognormal distribution.

The mean serial interval in Brazil of 2.97 days is the shortest reported to our knowledge, but we emphasize that the serial interval is not usually concentrated around the mean due to its high standard deviation (3.29 days) and non-symmetry. Furthermore, our serial interval estimate is shorter than the reported incubation period, implying that the infectious period for SARS-CoV-2 begins before symptom onset. This is in line with recent observations of a large proportion of undocumented infections, complicating containment of the virus [10]. The short mean observed, and the relatively high probability of zero or negative serial intervals reinforces previous results from Ganyani et al. [9], which estimate a large proportion of pre-symptomatic transmission for Singapore and Tianjin. Together, our findings imply that even with screening measures for travelers in airports, cruises or bus stations, the risk of transmission might remain relatively high.

Our report presents the first serial interval estimates for SARS-CoV-2 from Latin America. Our estimates are based on national case notification data, and this is a strength compared to datasets compiled from media reports. However, there are at least two important sources of bias. Firstly, there is a tendency for secondary cases to recall more recent contacts – recency or recall bias – resulting in a shorter estimate of the serial interval. Similarly, contact tracing is likely to be most effective for recent contacts. Secondly, self-isolation following symptom onset will remove longer serial intervals that would have occurred due to transmission during the symptomatic phase. This second point highlights the role of contextual factors such as isolation practices, population density, location of transmission, proportion of the population infected, in determining the serial interval.

In conclusion, we estimate a mean serial interval of COVID-19 at 2.97 days for Brazil. This estimate shorter than published incubation periods, implying pre-symptomatic undocumented transmission. Our estimate includes a substantial proportion of negative serial intervals that implies a substantial proportion of secondary transmission prior to the onset of symptoms.

## Data Availability

Data for this analysis were provided by the Brazilian Ministry of Health.

## Acknowledgements

We thank Philippe Mayaud and Oliver Brady for comments and suggestions in an earlier draft of this work. This work was supported by a Medical Research Council (MR/S0195/1) and FAPESP (2018/14389-0) CADDE partnership award and a John Fell Research Fund (grant 005166). NRF is supported by a Wellcome Trust and Royal Society Sir Henry Dale Fellowship (204311/Z/16/Z). DDSC is supported by the Clarendon Fund and by the Oxford University Zoology Department. CAPJ and VHN are supported in part by FAPESP grant 2018/12579-7 and CAPES. CAD is thankful for Centre funding from the UK Medical Research Council (MRC) and the UK Department for International Development (DFID) under the MRC/DFID Concordat agreement and is also part of the EDCTP2 programme supported by the European Union (MR/R015600/1). AD is also supported by the UK Medical Research Council (MRC).

